# Are quantitative radiomics features comparable to semantic radiology features for pre-operative risk classification of thymic epithelial tumours?

**DOI:** 10.1101/2025.04.26.25326466

**Authors:** Amal Joseph Varghese, Miracle Pathinathan, Balu Krishna Sasidharan, C Praveenraj, Rajendra Benny K, Thomas Kodiatte, Manu Mathew, I Rajesh, Simon Pavamani, Aparna Irodi, Leonard Wee, Andre Dekker, Hannah Mary Thomas T

## Abstract

Thymic epithelial tumours (TETs) are rare and exhibit varied behaviour and prognosis based on their histological subtype, as classified by the World Health Organization (WHO). These subtypes are further categorized into low-risk and high-risk groups. Low-risk thymomas generally allow for complete surgical resection without adjuvant therapy, while high-risk types often require multimodal treatment due to their aggressive nature. This study aims to evaluate the role of CT radiomics in discriminating between high- and low-risk TETs.

**Methods:** This retrospective study included patients treated in a single hospital in India who underwent surgical resection of TETs from 2010 to 2024. Inclusion criteria were confirmed TET diagnosis, had pre-operative CT scans, and had medical and post-operative histopathological confirmation. Conventional CT (semantic) features were manually extracted from radiology reports, while radiomic features were obtained using PyRadiomics. The data was randomly split for training and a hold-out validation set stratified by class. Three classification models were evaluated, each using clinical, semantic, and radiomic features with LASSO regularization. The performance of the models was assessed using Area under the Receiver Operating Curve (AUC), sensitivity, and specificity on the test set.

**Results:** Out of 195 enrolled patients, 132 met inclusion criteria and were divided into training (n = 100) and a validation set (n=32). The clinical model included age, presence of Pure Red Cell Aplasia and weight loss, achieving an AUC of 0.69 (95% CI: 0.49–0.87), sensitivity of 0.73 (95% CI: 0.46-1.00), and specificity of 0.53 (95% CI: 0.29-0.76) in the holdout set. The radiomics model included 90th percentile and sphericity as key predictors with AUC of 0.77 (95% CI: 0.56-0.94), sensitivity of 0.82 (95% CI: 0.55–1.00), and specificity of 0.72 (95% CI: 0.52-0.91). The semantic model performed best with AUC of 0.82 (95% CI: 0.62– 0.96), sensitivity of 0.82 (95% CI: 0.55-1.00), and specificity of 0.77 (95% CI: 0.57-0.91).

**Discussion and Conclusion:** The findings indicate that radiomic features could be valuable in preoperative risk assessment for TETs. Although the conventional CT features-based Semantic models demonstrated superior predictive capability, there is a risk of subjectivity and inter-observer disagreement.

## Introduction

With an incidence rate of only 1.5 cases per million people, Thymic Epithelial tumours (TETs) are rare neoplasms [1–3]. TETs comprise of thymomas, thymic carcinomas, and thymic neuroendocrine tumours and these all originate in the thymus gland. Thymomas are often found in the anterior mediastinum of adults. Thymic carcinomas are less common and are often present with metastasis, leading to a significantly worse prognosis compared to thymomas. Hence, TETs represent a heterogeneous group of tumours [2,4]. Despite favourable survival rates, it is the histological subtype that often significantly impacts the prognosis and response to treatment.

The World Health Organization (WHO) classifies thymic epithelial tumours as type A, AB, B1, B2, B3, or thymic carcinoma, on the basis of the morphology of their epithelial cells and the ratio of lymphocytes to epithelial cells. Types A, AB and B1 are collectively stratified as low risk, and types B2-B3 are defined as high risk. [5–7]. Low risk thymomas are often surgically resected without the need for adjuvant therapy, whereas the high-risk thymomas may require multimodal treatment because there is a higher chance of local invasion which reduces the possibility of a total resection. Thymic carcinomas are included in the high-risk category due to their aggressive nature and poor prognosis, and require a multi-modal therapeutic approach. Therefore, histological classification is crucial for risk stratification and tailoring surgical or radiation treatment for these patients.

The conventional method of surgical resection for thymoma is median sternotomy, which is gradually being replaced by minimally invasive robotic-assisted and video-assisted thoracoscopic surgery [8–10]. However, minimally invasive surgery for high-risk thymomas and thymic carcinomas is still controversial due to concerns over tumour manipulation, capsular disruption and partial resection that can eventually lead to local recurrence and reduced survival [11,12].

Hence, preoperatively ascertaining the TET’s risk grouping in a timely manner would allow the surgeons to choose the most appropriate surgical approach. Suitable serum markers are not widely available. However, contrast-enhanced computed tomography (CT) is almost ubiquitous and regularly used for radiological assessment of TETs[13–17].

However, the qualitative reporting of the CT for this rare type of tumour is not always reliable since the interpretation of imaging is subjective and depends on radiologist experience. The next best approach is a pre-surgical needle biopsy. However, this is invasive, and the biopsy sampling might be subject to tumour subsampling heterogenetiy and might not fully represent the entirety of a TET. Hence an efficient, low-cost and non-invasive preoperative technique would be valuable to guide surgical decision-making in managing TETs.

Radiomics is a growing research area that examines the links between quantitative data from medical images and clinical information. The key idea is that differences in imaging reflect distinct tumour phenotypes, revealing valuable prognostic information that is not visible to the naked eye. Unlike biopsies or molecular markers that examine only specific locations, radiomics assesses the entire tumour, minimizing sampling errors. This is especially relevant for managing thymic epithelial tumours, as understanding the variations within a tumour is as crucial as differences between the tumours for predicting the risk. A few recent studies have shown that Radiomics has promise in differentiating between the risk categories in Thymomas [13,14,16–18]. However, its prognostic role within a South Asian population has not been studied.

The aim of this study is to evaluate the role of CT radiomics in distinguishing between high- and low-risk TET groups to stratify them for pre-operatively based on their risk. The Radiomics Risk Model is compared to the clinical and radiologist reported qualitative CT features for this rare disease type in a South Asian population treated in a single institution.

## Methods and Materials

This ethics-approved (ref 14834) retrospective study included all patients treated for TETs, who underwent surgical resection between May 2010 and May 2024 at our hospital, and were all pathologically confirmed as being TETs. The inclusion criteria were as follows: Patients with diagnosed TETs and assigned WHO grading: 1) had pre-operative arterial phase contrast enhanced computed tomography of thorax 2) no treatment before CT scan; and (3) histopathological reports. The WHO grading was based on either surgical resection or small biopsy specimens only. The exclusion criteria were as follows 1) poor image quality due to artefacts or other causes. 2) Incomplete clinical records. The study cohort was selected based on the specified criteria as illustrated in Fig. 1. The ethical committee waived the need to re-ask the patients for informed consent to use their personal information due to the retrospective nature of the chart study based on routine-of-care data.

**Figure 1:**
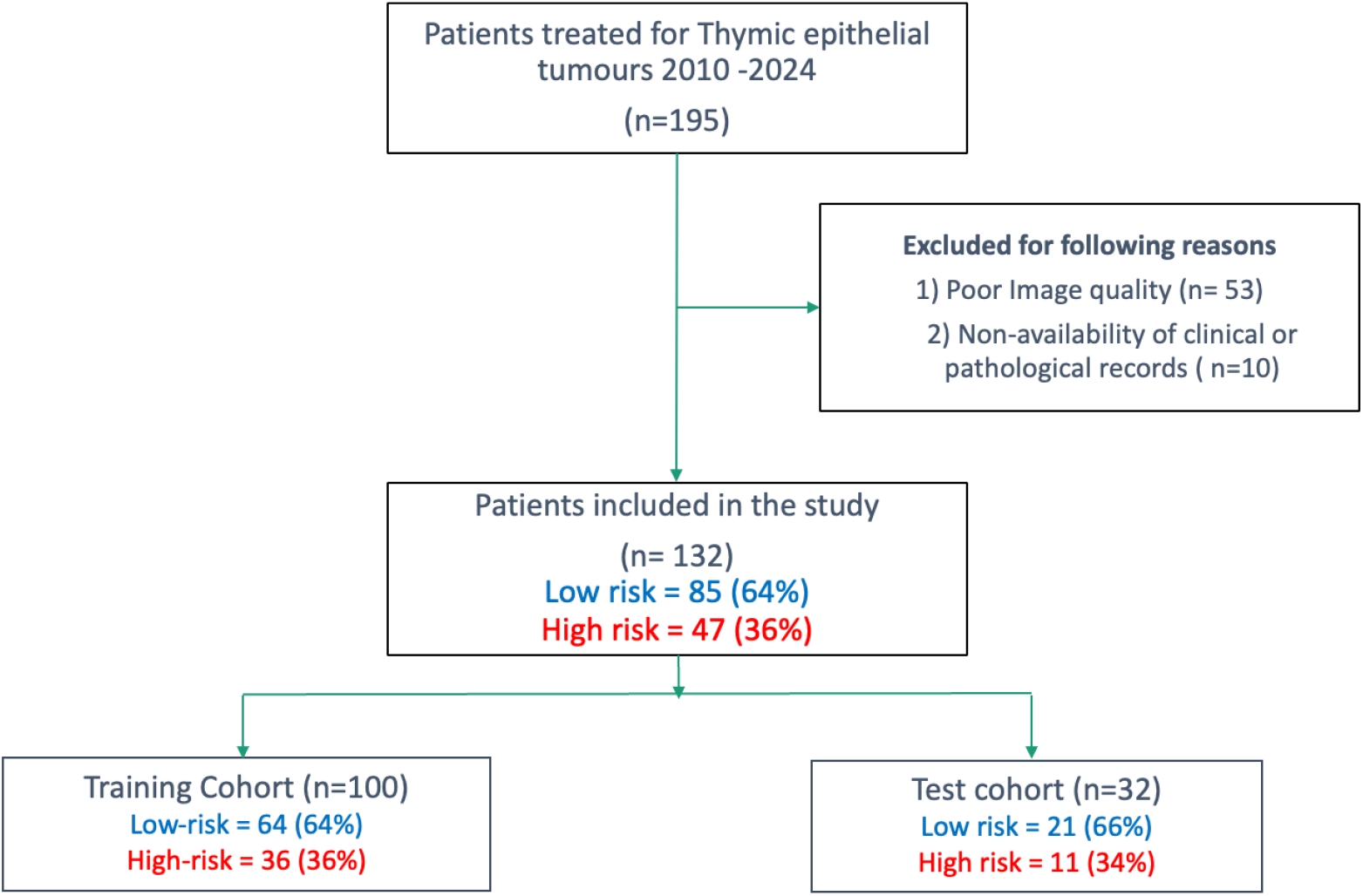
Cohort selection diagram

### Imaging

CT images were obtained using a 64 slice multi-detector CT scanner (Discovery CT 750 HD, GE Healthcare, Waukesha, WI, United States). The scan parameters included for automatic modulation were 120 kV tube voltage; tube rotation time 0.6s, pitch 0.531:1, slice thickness of 2.5 or 5 mm; reconstruction interval of 2.5 mm. Non-ionic contrast material (Iohexol 300) was injected at 3 mL/s using an IV cannula (20G) with a mechanical automated pressure injector. The CE-CT images of arterial phase were collected 40 seconds following the injection using bolus tracking. Images were acquired from the chin to the lower border of liver and viewed with standard mediastinal (level -80 HU, width-300 HU) and lung (level-500 HU; width-1500 HU) and narrower liver (level-88 HU, width-150 HU) windows.

### Conventional CT Features

The conventional CT image findings commonly identified and rated by radiologists based on their expertise and visual assessment were manually extracted from the radiology reports. There were 25 features that were included -which we call “semantic features”; descriptors of the tumour contour (i.e. smooth, lobulated or irregular), tumour margin, tumour shape (i.e. rounded, oval, irregular), side and location of the tumour (upper third, mid third, lower third, upper & mid third, mid & lower third, upper-mid-lower third), homogeneity of enhancement (homogenous or heterogeneous), degree of enhancement (i.e. hypo, iso or hyper enhancing), enhancement ratio, maximum tumour size (measured as the largest cross-section of the mass), the presence of necrosis, cystic degeneration, central contrast pooling, nodular enhancement, calcification, satellite lesions, mediastinal fat infiltration, fat stranding, fat plane obliteration with lung or CVS, mediastinal lymphadenopathy, irregular infiltration of pleura, pleural deposits and pleural effusion, presence of distant metastasis and hypertrophied vessels.

### Radiomics ROI segmentation

The regions of interest (ROI) were delineated on the CE-CTs using the delineation tools available on the Varian (Palo Alto, CA, USA) Eclipse treatment planning system. The ROI included the edges of the lesion identified as the TET, including calcification. ROIs were manually segmented first by a general radiologist (MP) and reviewed independently by a senior thoracic radiologist (AI), and finally by a senior radiation oncologist (BS) who approved or corrected the segmentation. Any discrepancies were resolved by discussion between these three clinicians until a consensus was reached.

### Radiomics Feature extraction and selection

The radiomic “original” features **(n=103)** as referred to by Aerts et al.[19] were extracted from each TET ROI using PyRadiomics (v3.0.1), which is mostly compliant with the Image Biomarker Standardization Initiative (IBSI); deviations – if any -were stated in the developer’s documentation and were not within control of the study authors. The extracted features were divided into six classes; first order features, shape features, gray level co-occurrence matrix-based features (GLCM), gray level run length matrix-based features (GLRLM), gray level size zone matrix-based features (GLSZM), gray level dependence matrix-based features (GLDM), and neighborhood gray tone difference matrix-based features (NGTDM). Post-filter features such as LoG or Wavelet filtered features were excluded. Based on the training dataset each extracted radiomics feature was normalized by centring the mean to 0 and scaled to sample standard deviation of 1.

Before feature selection, the data was randomly split into training (76%) and testing (hold-out) sets (24%), stratified by class. Feature selection was conducted only on the training data in a multi-step process: 1) Features highly correlated with each other or with tumor size were removed based on a Pearson correlation. Only features with a correlation coefficient below the set threshold of 0.75 were retained. 2) SelectKBest (from the Python scikit-learn library) was used to rank the features based on their statistical relationship with the target outcome, using one-way ANOVA as the scoring function. 3) Logistic Regression models with LASSO regularization were used.

The number of features to be retained via SelectKBest (k) and the strength of the regularization parameter (C) were optimized using 5-fold cross-validation and default grid search, with ROC-AUC as the scoring method. The values for ‘k’ tested were between 1 to 15 and C was tested using 10 values on a logarithmic scale between 1e^-4^ and 1e^4^. The features producing the best performance after 1000 iterations were selected.

### Model construction and Testing

Using the features and model parameters selected by the 5-fold grid search cross-validation and based on the Radiomic features, we derived the rad-scores from the logit functions that were used to generate the probability metric [20]. Similarly, clinical and semantic scores were generated using the selected features from their respective models. These scores were used to create combined models such as clinical-semantic (CS-model), semantic-radiomics (SR-model), clinical-radiomics (CR-model) and clinical-semantic-radiomics (CSR-model).

All models were fit on the entire training data set and repeated the same steps in the hold-out validation data set without any re-computation. area under the receiver operating-curve (AUC), accuracy, specificity and sensitivity were used to evaluate the performance of the models and were reported over 1000 bootstrap. A risk threshold of 0.36, equal to the true event fraction of training dataset was used to calculate the accuracy, sensitivity and specificity. This threshold was selected based on the work of van den Goorbergh et al to mitigate the class imbalance present in training data [21].

### Statistical Analysis

Patient and clinical characteristics were compared between patients with high and low-risk WHO grades, and between training and testing sets, using Fisher’s exact test, and the Chi-square test and t-test, where appropriate P-values < 0.05 were considered statistically significant. Machine learning and statistical analyses were performed using the Sci-kit library and Python libraries (Version 3.7.13). Graphs were generated using Matplot and Seaborn libraries.

## Results

A total of 195 patients were enrolled in our study, and 132 patients were included for final analysis. Table 1 lists patient characteristics. The low-risk group included 85 patients (type A: 7, type AB: 51, type B1: 27), and the high-risk group included 47 patients (type B2: 34, type B3: 10 and thymic carcinoma: 3). No significant differences were observed between the low- and high-risk groups in either the training cohort (n = 100 cases) or the test cohort (n =32) (Table 1).

**Table 1:**
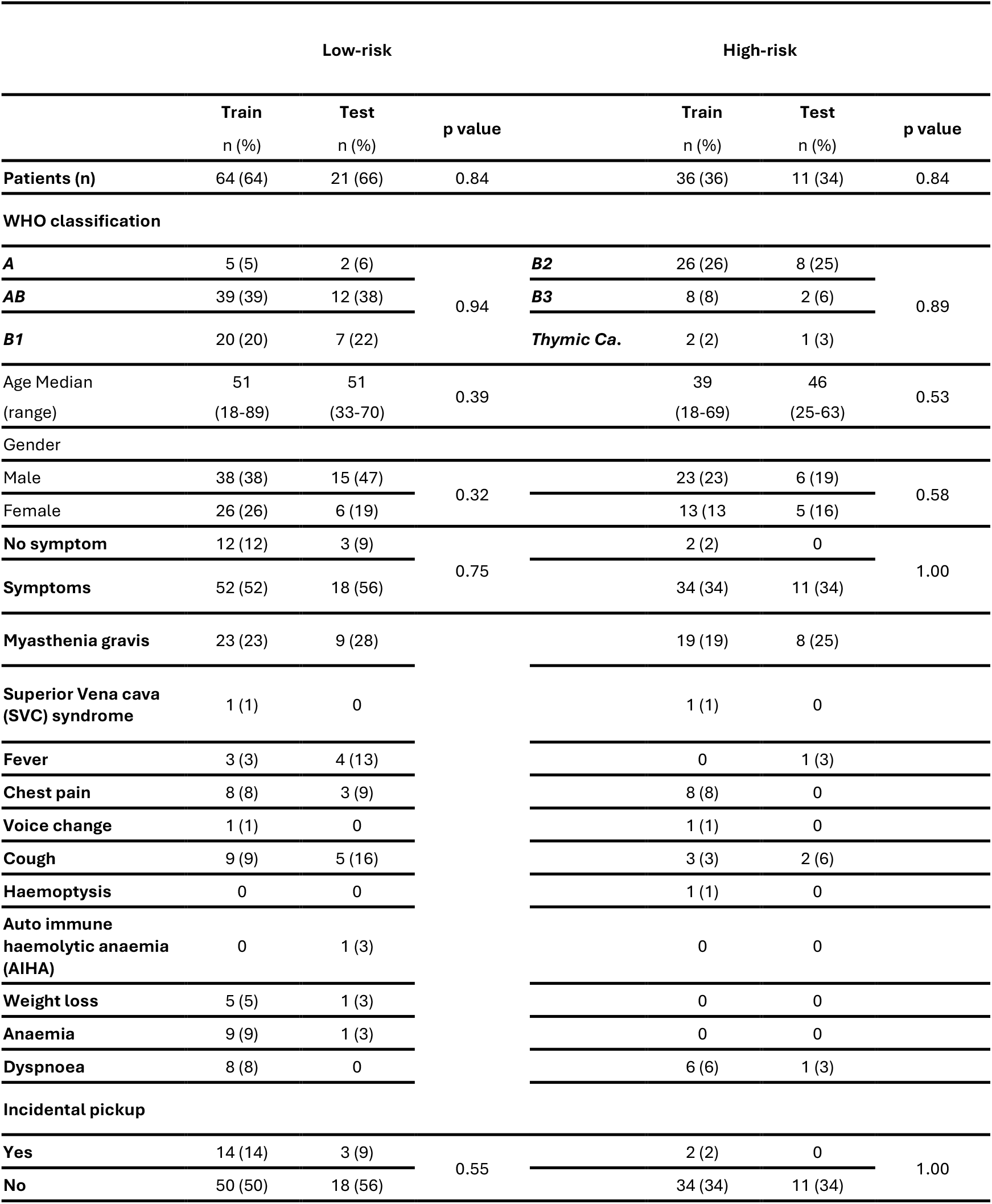
Patient characteristics.

Figure 2 presents the ROC curve analysis results for the training and test datasets to differentiate between the low-risk and high-risk groups of TETs.

**Figure 2:**
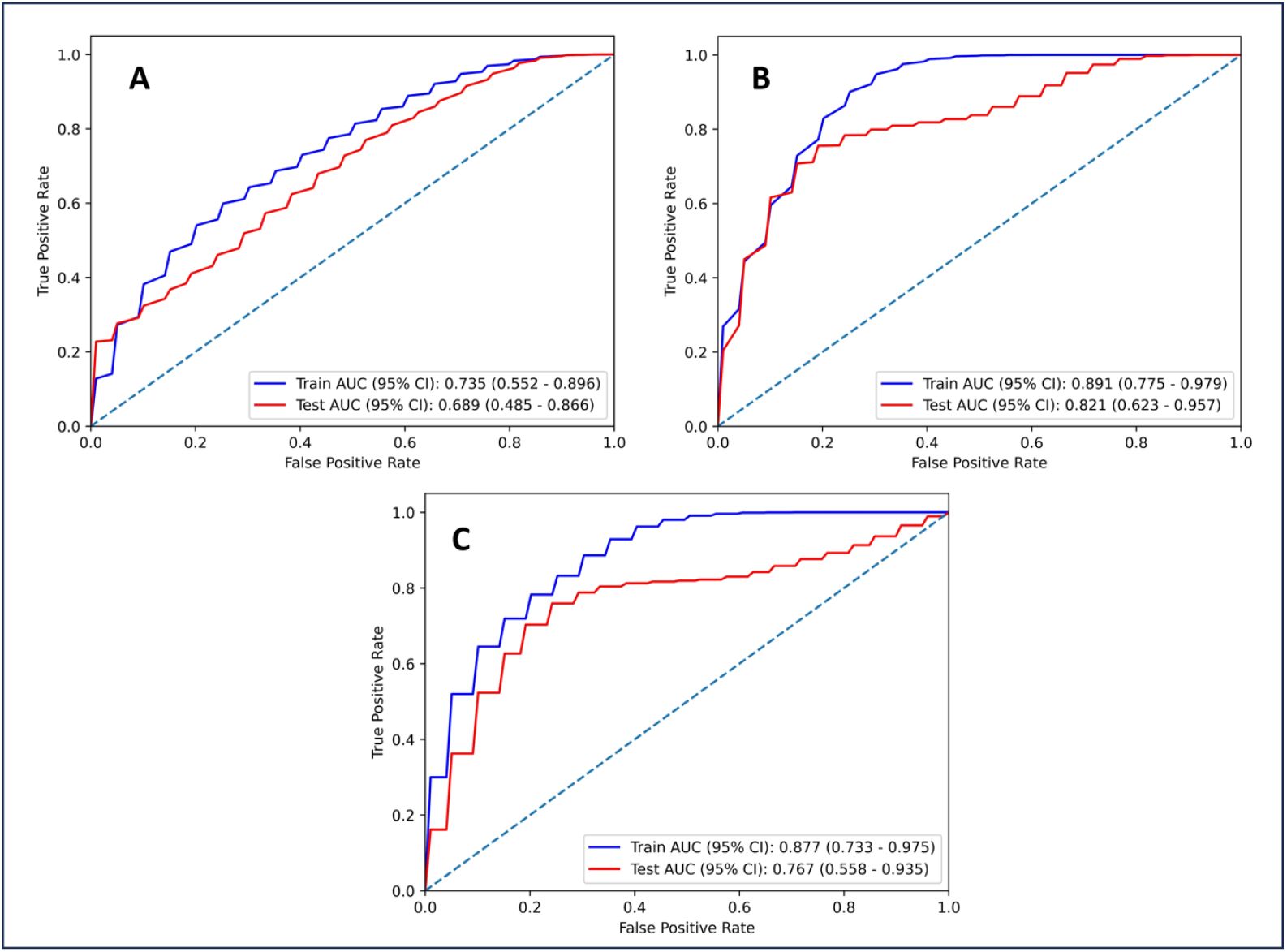
Performance of A. Clinical B. Semantic C. Radiomics models as represented by the Area under the Receiver Operating Curves in the training (blue) and hold-out test (red) datasets.

Table 2 represents the performance of the models created using the Clinical, Semantic and Radiomics features independently. The Semantic model included tumour shape, tumour contour, tumour margin, tumour location, areas of nodular enhancement and degree of enhancement as the key predictors for risk stratification of TETs and performed the best with a test AUC of 0.82 (95% CI: 0.62–0.96) and sensitivity of 0.82 (95% CI: 0.55-1.00) and specificity of 0.77 (95% CI:0.57-0.91). The Radiomics model included two main features namely, 90^th^percentile and sphericity and achieved an AUC of 0.77 (95% CI: 0.56–0.94), with a sensitivity of 0.82 (95% CI: 0.55–1.00) and a specificity of 0.72 (95% CI: 0.52-0.91). The clinical RM included three features namely age, weight loss and presence

**Table 2:** Performance results of the individual & combined models.

| Model | Task | AUC<br>(95 % CI) | Accuracy<br>(95 % CI) | Sensitivity<br>(95 % CI) | Specificity<br>(95 % CI) |
| --- | --- | --- | --- | --- | --- |
| **Clinical** | Training | 0.74<br>(0.55 - 0.90) | 0.69<br>(0.56 - 0.84) | 0.75<br>(0.50 - 1.00) | 0.58<br>(0.35 - 0.80) |
|  | Test | 0.69<br>(0.49 - 0.87) | 0.69<br>(0.59 - 0.78) | 0.73<br>(0.46 - 1.00) | 0.53<br>(0.29 - 0.76) |
| **Semantic** | Training | 0.89<br>(0.78 - 0.979) | 0.80<br>(0.66 - 0.91) | 0.92<br>(0.75 - 1.00) | 0.75<br>(0.55 - 0.90) |
|  | Test | 0.82<br>(0.62 - 0.96) | 0.85<br>(0.719 - 0.97) | 0.82<br>(0.55 - 1.00) | 0.77<br>(0.57 - 0.91) |
| **Radiomics** | Training | 0.88<br>(0.73 - 0.98) | 0.78<br>(0.63 - 0.91) | 0.89<br>(0.67 - 1.00) | 0.68<br>(0.45 - 0.85) |
|  | Test | 0.77<br>(0.56 - 0.94) | 0.78<br>(0.63 - 0.91) | 0.82<br>(0.55 - 1.00) | 0.72<br>(0.52 - 0.91) |
| **Clinical-Semantic** | Training | 0.92<br>(0.80 - 1.00) | 0.79<br>(0.66 - 0.91) | 0.92<br>(0.75 - 1.00) | 0.85<br>(0.70 - 1.00) |
|  | Test | 0.86<br>(0.71 - 0.97) | 0.79<br>(0.66 - 0.91) | 0.82<br>(0.55 - 1.00) | 0.81<br>(0.62 - 0.95) |
| **Semantic-Radiomics** | Training | 0.91<br>(0.79 - 0.98) | 0.78<br>(0.63 - 0.91) | 0.92<br>(0.75 - 1.00) | 0.78<br>(0.60 - 0.95) |
|  | Test | 0.86<br>(0.70 - 0.98) | 0.78<br>(0.63 - 0.91) | 0.81<br>(0.55 - 1.00) | 0.71<br>(0.52 - 0.91) |
| **Clinical-Radiomics** | Training | 0.90<br>(0.77 - 0.98) | 0.82<br>(0.69 - 0.94) | 0.81<br>(0.58 - 1.00) | 0.80<br>(0.60 - 0.95) |
|  | Test | 0.79<br>(0.59 - 0.95) | 0.79<br>(0.63 - 0.94) | 0.83<br>(0.55 - 1.00) | 0.75<br>(0.57 - 0.95) |
| **Clinical-Semantic-Radiomics** | Training | 0.93<br>(0.82 - 1.00) | 0.87<br>(0.75 - 0.97) | 0.98<br>(0.83 - 1.00) | 0.82<br>(0.60 - 0.95) |
|  | Test | 0.88<br>(0.73 - 0.98) | 0.78<br>(0.62 - 0.91) | 0.82<br>(0.55 - 1.00) | 0.71<br>(0.52 - 0.91) |

of anaemia to achieve a test AUC of 0.69 (95% CI: 0.49–0.87), sensitivity of 0.73 (95% CI: 0.46-0.1.00) and specificity of 0.53 (95% CI:0.29-0.76).

The equations for the clinical, sematic and rad-scores as derived from the individual RMs is shown in Supplementary A1. Figure 3 shows the ROC curve analysis results for the training and test datasets for the combined models. The performance metrics for the combined models is shown in supplementary Table A1.

**Figure 3:**
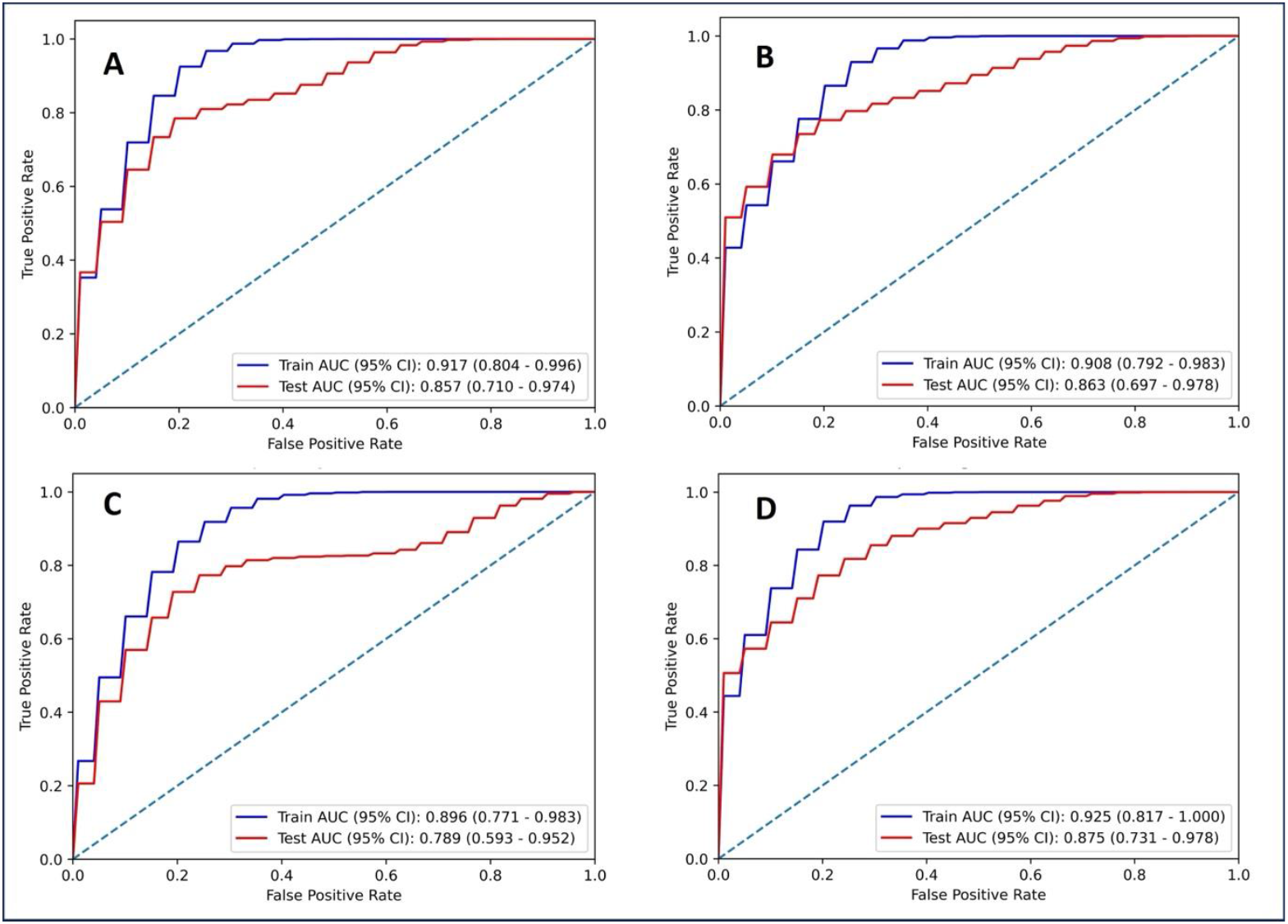
Performance of the combined models A. (Clinical-Semantic) B. (Semantic-Radiomics) C. (Clinical-Radiomics) D. (Clinical-Semantic-Radiomics) models as represented by the Area under the Receiver Operating Curves in the training (Blue) and in the hold-out test (Red) datasets.

## Discussion

This study developed risk models based on clinical, radiologist defined semantic descriptors and radiomics from CE-CT images using logistic regression models to predict the WHO classification of high and low risk groups in thymic epithelial tumours. A combined model incorporating the risk scores from clinical, semantic and radiomics models performed the best for the given dataset. To our knowledge, this is the first study investigating the role of Radiomics for risk stratification for TETs in a South Asian Population [22]. In clinical practice, accurately distinguishing among low-risk thymoma, high-risk thymoma, and thymic carcinoma is still difficult [18]. Most of thymic epithelial tumours are asymptomatic whereas those with symptomatic thymoma have advanced stage at diagnosis. Since thymoma is associated with myasthenia gravis, some of them may be diagnosed when imaging is done to look for the cause of myasthenia. However, the association between myasthenia gravis and thymoma is debatable [23,24]. In certain rare occasions, thymoma are diagnosed while evaluating refractory anaemia [25]. Our clinical model included ‘age’ as reported by Liu et al as a predictor of risk, along with the ‘weight loss’ and presence of ‘pure red cell aplasia’ (PRCA). Liu also reported that performance of the clinical model that included Age only improved when combined with radiomics [26], contrary to the results seen in our Clinical-Radiomics model. Previous studies have also attempted to differentiate low-risk thymoma, high-risk thymoma and thymic carcinoma based on the conventional CT findings (Semantics models) and have reported differences in tumour size and shape, adjacent mediastinal fat infiltration, invasion of large vessels between low-risk thymoma, high-risk thymoma, and thymic carcinoma [27–29]. However, there may be some lack of specificity in these findings and the diagnosis is highly dependent on the reporting radiologist’s experience [30].

Studies have shown that radiomics shows promise in differentiating between the risk groups [16–18,31–33] and our study concurs with these results. Our radiomics risk model included two main features namely, 90^th^ percentile and sphericity. Following the suggestion by Mattea Welch et al., we verified that these features were not a confounder for tumour volume (Supplementary Figure A1)[34]. Notably, fewer spherical tumours were seen in the high-risk class (Figure 4). The shape of the tumour has also been reported as a strong predictor for differentiating between high and low risk models by other studies [15,13].

**Figure 4:**
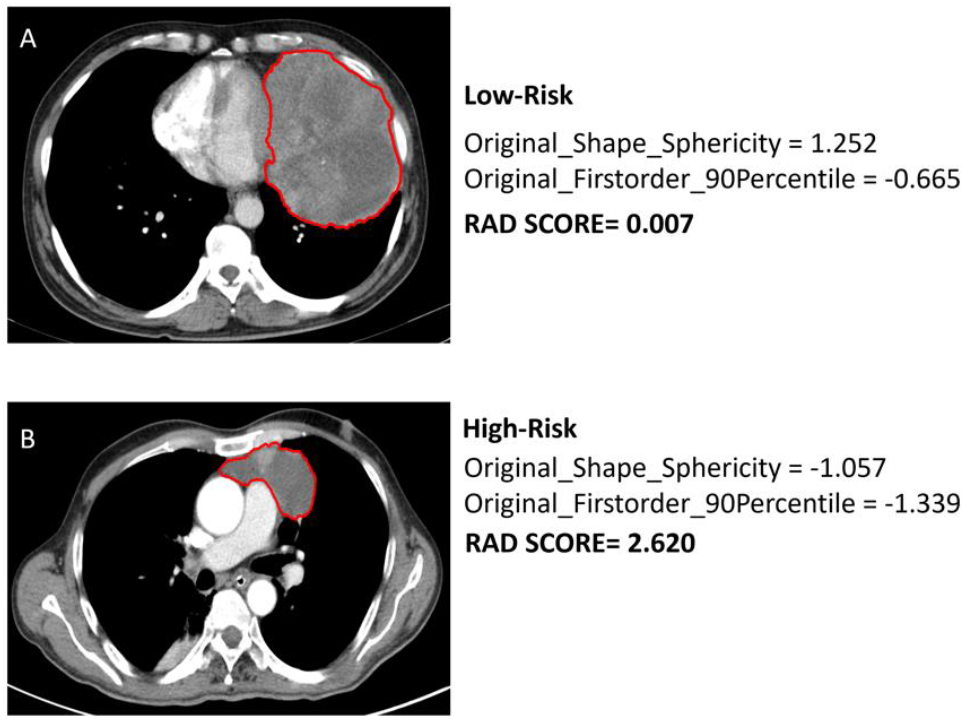
Representative patients with (A) Low and (B) high-risk TETs and their Radiomics feature values and rad-score.

Our semantic model displayed that the high-risk TETs displayed irregular shape and hyper-enhancing tumours without smooth contours. Interestingly, there was a high clustering between radiomics and semantic shape features (See Figure 5). Although our semantic model demonstrated superior predictive capability compared to the radiomics model in our dataset, it is qualitative and highly reliant on observer judgement. However, since the performance between the semantic and radiomics models was not too substantial, using the radiomics approach could improve decision making due to its objectivity and automatability. low risk. C and D show the WHO grade spread for the same. X and Y axis represent the standard deviation of the Radiomics features of the training set.

**Figure 5:**
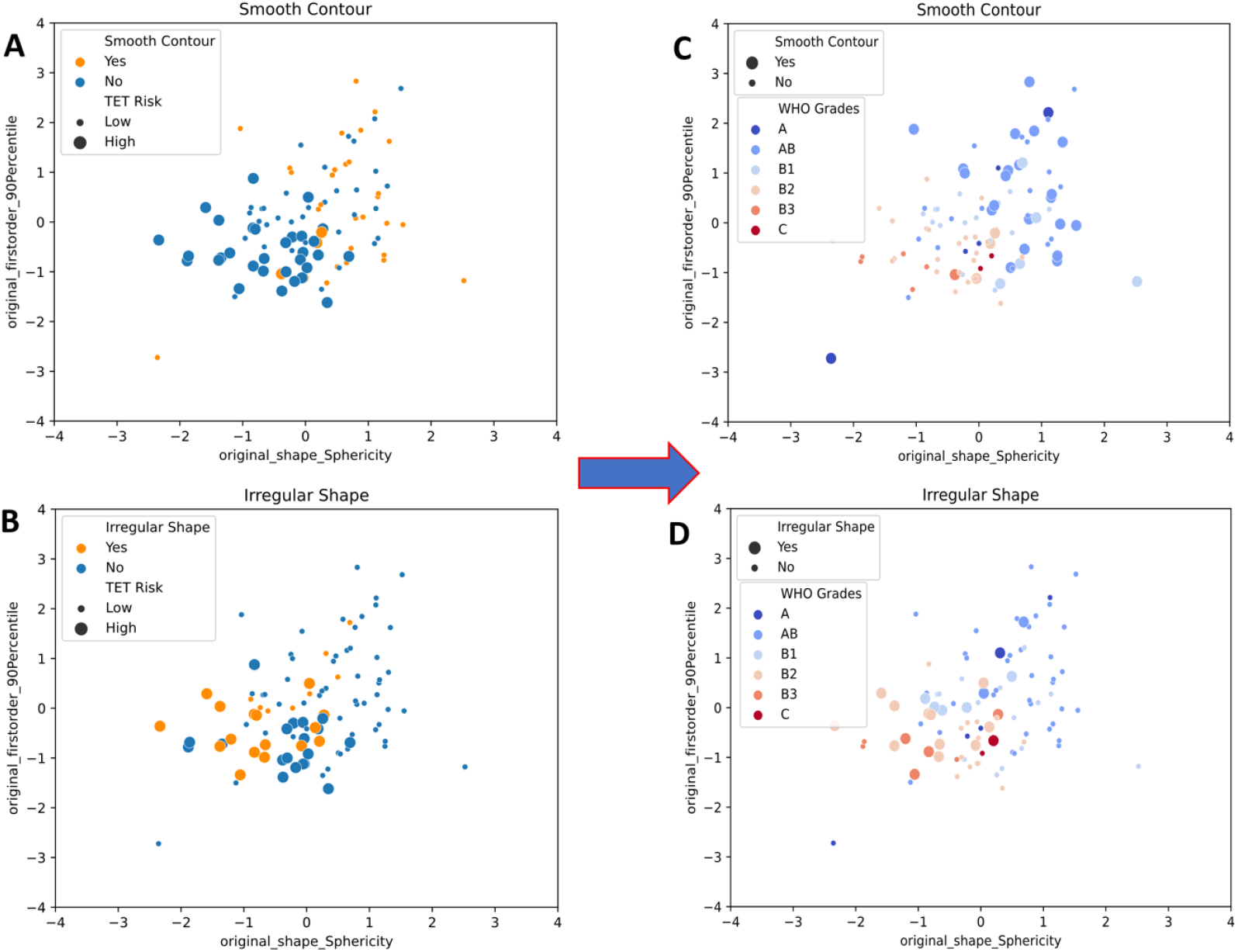
Relationship between the semantic shape features (Smooth contour and Irregular shape) and Radiomic features; A and B show the spread between the high and

Liu et al reported RMs to differentiate between low-risk (A, AB, B1), high-risk (B2, B3) and thymic carcinomas[18]. Others have included RMs only for Low-risk and high-risk as categorized by WHO and reported in [13,14,35]. However, Liang et al have included the thymic cancers to the high-risk category like our study [16]. This was done because the numbers for the thymic cancers were very low and the management follows a similar pattern to that of grades B2 and B3.

Many different Machine learning classifiers have been explored to find the best RMs for classifying high and low-risk TETs. Cangir et al explored 6 classifiers for their risk models namely k-Nearest Neighbour (KNN), Support Vector Machine (SVM), eXtreme Gradient Boosting (XGBoost), Random Forest (RF), Logistic Regression (LR), and Decision Tree (DT). and reported LR model to be the best classifier [14]. Similarly, other studies have also shown LR to perform well for CT-based radiomics to predict different risk subgroups of TETs or thymoma studies [17,26] and has influenced our choice to use LR algorithm for this study.

In our study, the semantic model (AUC 0.82) outperformed both clinical (AUC: 0,69) and radiomics (AUC: 0.77) models. Similar to other studies, we observed that the combined models generally outperform the individual models [13,16,35,36]. The semantics-radiomics and the clinical-semantics were roughly the same in overall discriminative performance AUC 0.86 (95 % CI 0.70-0.98) but the clinical-semantic-radiomics model performed the best with AUC 0.88 (95% CI: 0.73–0.98), sensitivity of 0.82 (95% CI: 0.55-0.1.00) and specificity of 0.71 (95% CI:0.52-0.91). However, in a data-driven approach, we have included the features as selected by the model. For example, ‘presence of PRCA’ was chosen as an important clinical feature but we cannot provide any direct insight into the biological meaning of the finding [33]. Our radiologists would not want to rely on this feature for predicting risk type of TETs. Hence, further study is required to see how to improve feature selection while considering data-driven approaches.

This study has a few limitations. First, it was a retrospective single centre study which makes selection bias inevitable. Second, our sample size was small but given TETs is a rare disease, this is a large number from a single institution. However, it should be noted that follow-up for these patients has been challenging. A multicentre study with a larger sample size will be required to ensure that the results are generalizable. Third, we did not specifically analyse each WHO pathological subtype individually within the semantic and radiomics models. Instead, we categorized the subtypes into low- and high-risk groups. Future studies can explore this further to enable prediction of specific pathological subtypes in the pre-operative setting Their importance should not be ignored and we hope future studies can combine them to predict the actual pathological type of the tumour and not limit it to the just risk category prediction. Next, overfitting is an important problem in machine learning when dealing with high-dimensional features with small sample size. In our study we tried to limit to 103 radiomics features instead of 1000+ features given our sample size was 131. We also attempted to mitigate overfitting by using 5-fold cross-validation and bootstrapping techniques and having a hold-out test set for evaluation of the results.

In conclusion, this study shows that radiomics shows promise as a pre-operative risk prediction tool for thymic epithelial tumours that can complement radiologists’ findings of the tumour. Complementary information from radiomics and clinical findings might help radiologist improve the pre-operative imaging-based risk stratification for thymic epithelial tumours. Prospective and larger studies involving more diverse populations are needed to validate these findings for clinical implementation of this this type of risk stratification.

## Data Availability

All data produced in the present study are available upon reasonable request to the authors.

## Acknowledgments

Authors AJV and HMT acknowledge the support of the DBT/Wellcome Trust India Alliance Early Career Fellowship [Grant number: IA/E/18/1/504306] awarded to HMT. Author BS acknowledges the support by the Foundation I-DAIR. Authors from the Department of Radiation Oncology, CMC Vellore acknowledge the support of the Tamil Nadu Dr. MGR Medical University.

## Notes

### Competing Interest Statement

The authors have declared no competing interest.

### Funding Statement

This study was supported by the Internal Fluid Research grant from the institution.

### Author Declarations

Ethical committee of Christian Medical College has approved this study and has waived the need for informed consent due to the retrospective nature of the study. All images were de-identified for analysis purposes.

## References

[1] Rathod S, Munshi A, Paul S, Ganesh B, Prabhash K, Agarwal JP. Thymoma: First large Indian experience. Indian J Cancer 2014;51:109. 10.4103/0019-509X.138144.

[2] Bernard C, Frih H, Pasquet F, Kerever S, Jamilloux Y, Tronc F, et al. Thymoma associated with autoimmune diseases: 85 cases and literature review. Autoimmun Rev 2016;15:82–92. 10.1016/j.autrev.2015.09.005.

[3] de Jong WK, Blaauwgeers JLG, Schaapveld M, Timens W, Klinkenberg TJ, Groen HJM. Thymic epithelial tumours: a population-based study of the incidence, diagnostic procedures and therapy. Eur J Cancer Oxf Engl 1990 2008;44:123–30. 10.1016/j.ejca.2007.11.004.

[4] Ried M, Marx A, Götz A, Hamer O, Schalke B, Hofmann H-S. State of the art: diagnostic tools and innovative therapies for treatment of advanced thymoma and thymic carcinoma. Eur J Cardio-Thorac Surg Off J Eur Assoc Cardio-Thorac Surg 2016;49:1545–52. 10.1093/ejcts/ezv426.

[5] Guidelines Detail. NCCN n.d. https://www.nccn.org/guidelines/guidelines-detail?category=1&id=1469 (accessed November 6, 2024).

[6] Ruffini E, Detterbeck F, Van Raemdonck D, Rocco G, Thomas P, Weder W, et al. Tumours of the thymus: a cohort study of prognostic factors from the European Society of Thoracic Surgeons database. Eur J Cardiothorac Surg 2014;46:361–8. 10.1093/ejcts/ezt649.

[7] Suster S. The WHO 2021 thymoma classification: a work in progress. J Cancer Metastasis Treat 2022;8:N/A-N/A. 10.20517/2394-4722.2021.203.

[8] Yang Y, Dong J, Huang Y. Thoracoscopic thymectomy versus open thymectomy for the treatment of thymoma: A meta-analysis. Eur J Surg Oncol J Eur Soc Surg Oncol Br Assoc Surg Oncol 2016;42:1720–8. 10.1016/j.ejso.2016.03.029.

[9] O’Sullivan KE, Kreaden US, Hebert AE, Eaton D, Redmond KC. A systematic review of robotic versus open and video assisted thoracoscopic surgery (VATS) approaches for thymectomy. Ann Cardiothorac Surg 2019;8:17493–17193. 10.21037/acs.2019.02.04.

[10] Zhu L, Zhang L, Zuo C, Sun T, Jiang B. Robot versus video-assisted thoracoscopic thymectomy for large thymic epithelial tumors: a propensity-matched analysis. BMC Surg 2023;23:330. 10.1186/s12893-023-02228-8.

[11] Zhang Y, Lin D, Aramini B, Yang F, Chen X, Wang X, et al. Thymoma and Thymic Carcinoma: Surgical Resection and Multidisciplinary Treatment. Cancers 2023;15:1953. 10.3390/cancers15071953.

[12] Friedant AJ, Handorf EA, Su S, Scott WJ. Minimally invasive versus open thymectomy for thymic malignancies: systematic review and meta-analysis. J Thorac Oncol Off Publ Int Assoc Study Lung Cancer 2016;11:30. 10.1016/j.jtho.2015.08.004.

[13] Gao C, Yang L, Xu Y, Wang T, Ding H, Gao X, et al. Differentiating low-risk thymomas from high-risk thymomas: preoperative radiomics nomogram based on contrast enhanced CT to minimize unnecessary invasive thoracotomy. BMC Med Imaging 2024;24:197. 10.1186/s12880-024-01367-5.

[14] Kayi Cangir A, Orhan K, Kahya Y, Özakıncı H, Kazak BB, Konuk Balcı BM, et al. CT imaging-based machine learning model: a potential modality for predicting low-risk and high-risk groups of thymoma: “Impact of surgical modality choice.” World J Surg Oncol 2021;19:147. 10.1186/s12957-021-02259-6.

[15] Wang X, Sun W, Liang H, Mao X, Lu Z. Radiomics Signatures of Computed Tomography Imaging for Predicting Risk Categorization and Clinical Stage of Thymomas. BioMed Res Int 2019;2019:3616852. 10.1155/2019/3616852.

[16] Liang Z, Li J, Tang Y, Zhang Y, Chen C, Li S, et al. Predicting the risk category of thymoma with machine learning-based computed tomography radiomics signatures and their between-imaging phase differences. Sci Rep 2024;14:19215. 10.1038/s41598-024-69735-3.

[17] Dong W, Xiong S, Lei P, Wang X, Liu H, Liu Y, et al. Application of a combined radiomics nomogram based on CE-CT in the preoperative prediction of thymomas risk categorization. Front Oncol 2022;12:944005. 10.3389/fonc.2022.944005.

[18] Liu W, Wang W, Guo R, Zhang H, Guo M. Deep learning for risk stratification of thymoma pathological subtypes based on preoperative CT images. BMC Cancer 2024;24:651. 10.1186/s12885-024-12394-4.

[19] Aerts HJWL, Velazquez ER, Leijenaar RTH, Parmar C, Grossmann P, Carvalho S, et al. Decoding tumour phenotype by noninvasive imaging using a quantitative radiomics approach. Nat Commun 2014;5:1–9. 10.1038/ncomms5006.

[20] Zheng B-H, Liu L-Z, Zhang Z-Z, Shi J-Y, Dong L-Q, Tian L-Y, et al. Radiomics score: a potential prognostic imaging feature for postoperative survival of solitary HCC patients. BMC Cancer 2018;18:1148. 10.1186/s12885-018-5024-z.

[21] van den Goorbergh R, van Smeden M, Timmerman D, Van Calster B. The harm of class imbalance corrections for risk prediction models: illustration and simulation using logistic regression. J Am Med Inform Assoc JAMIA 2022;29:1525–34. 10.1093/jamia/ocac093.

[22] Lu X-F, Zhu T-Y. Diagnostic performance of radiomics model for preoperative risk categorization in thymic epithelial tumors: a systematic review and meta-analysis. BMC Med Imaging 2023;23:115. 10.1186/s12880-023-01083-6.

[23] Cacho-Díaz B, Salmerón-Moreno K, Lorenzana-Mendoza NA, Texcocano J, Arrieta O. Myasthenia gravis as a prognostic marker in patients with thymoma. J Thorac Dis 2018;10. 10.21037/jtd.2018.04.95.

[24] Kondo K, Monden Y. Thymoma and Myasthenia Gravis: A Clinical Study of 1,089 Patients From Japan. Ann Thorac Surg 2005;79:219–24. 10.1016/j.athoracsur.2004.06.090.

[25] Gendron N, Fontbrune FS de, Guyard A, Fadlallah J, Chantepie S, D’Aveni M, et al. Aplastic anemia related to thymoma: a survey on behalf of the French reference center of aplastic anemia and a review of the literature. Haematologica 2020;105:e333. 10.3324/haematol.2019.226134.

[26] Liu J, Yin P, Wang S, Liu T, Sun C, Hong N. CT-Based Radiomics Signatures for Predicting the Risk Categorization of Thymic Epithelial Tumors. Front Oncol 2021;11.

[27] Strange CD, Ahuja J, Shroff GS, Truong MT, Marom EM. Imaging Evaluation of Thymoma and Thymic Carcinoma. Front Oncol 2022;11:810419. 10.3389/fonc.2021.810419.

[28] Jeong YJ, Lee KS, Kim J, Shim YM, Han J, Kwon OJ. Does CT of Thymic Epithelial Tumors Enable Us to Differentiate Histologic Subtypes and Predict Prognosis? Am J Roentgenol 2004;183:283–9. 10.2214/ajr.183.2.1830283.

[29] Gentili F, Monteleone I, Mazzei FG, Luzzi L, Roscio DD, Guerrini S, et al. Advancement in Diagnostic Imaging of Thymic Tumors. Cancers 2021;13:3599. 10.3390/cancers13143599.

[30] Marom EM. Imaging Thymoma. J Thorac Oncol 2010;5:S296–303. 10.1097/JTO.0b013e3181f209ca.

[31] Ohira R, Yanagawa M, Suzuki Y, Hata A, Miyata T, Kikuchi N, et al. CT-based radiomics analysis for differentiation between thymoma and thymic carcinoma. J Thorac Dis 2022;14:1342–52. 10.21037/jtd-21-1948.

[32] Iannarelli A, Sacconi B, Tomei F, Anile M, Longo F, Bezzi M, et al. Analysis of CT features and quantitative texture analysis in patients with thymic tumors: correlation with grading and staging. Radiol Med (Torino) 2018;123:345–50. 10.1007/s11547-017-0845-4.

[33] Yasaka K, Akai H, Nojima M, Shinozaki-Ushiku A, Fukayama M, Nakajima J, et al. Quantitative computed tomography texture analysis for estimating histological subtypes of thymic epithelial tumors. Eur J Radiol 2017;92:84–92. 10.1016/j.ejrad.2017.04.017.

[34] Welch ML, McIntosh C, Haibe-Kains B, Milosevic MF, Wee L, Dekker A, et al. Vulnerabilities of radiomic signature development: The need for safeguards. Radiother Oncol 2019;130:2–9. 10.1016/j.radonc.2018.10.027.

[35] Shang L, Wang F, Gao Y, Zhou C, Wang J, Chen X, et al. Machine-learning classifiers based on non-enhanced computed tomography radiomics to differentiate anterior mediastinal cysts from thymomas and low-risk from high-risk thymomas: A multi-center study. Front Oncol 2022;12. 10.3389/fonc.2022.1043163.

[36] Shen Q, Shan Y, Xu W, Hu G, Chen W, Feng Z, et al. Risk stratification of thymic epithelial tumors by using a nomogram combined with radiomic features and TNM staging. Eur Radiol 2021;31:423–35. 10.1007/s00330-020-07100-4.

[37] Tomaszewski MR, Gillies RJ. The Biological Meaning of Radiomic Features. Radiology 2021;298:505–16. 10.1148/radiol.2021202553.

