## Supplementary file for "Are quantitative radiomics features comparable to semantic radiology features for pre-operative risk classification of thymic epithelial tumours?"

### Supplementary files

### Section A1. Score Calculation

**Rad Score** = 0.613 + (-0.902 × original_shape_Sphericity) + (-0.787 × original_firstorder_90Percentile)

**Semantic Score** = -0.393 + (-2.195 × Tumour Contour) + (0.01 × Tumour Margin) + (1.566 × Tumour Shape) + (0.611 × Tumour Location) + (2.213 × Hypoenhancing Attenuation) + (-1.005 × Hyperenhancing Attenuation) + (-1.687 × Nodular Enhancement)

**Clinical Score** = 1.503 + (-4.134 × Age) + (-3.806 × Weight Loss) + (-4.537 × PRCA) s

Note: original_shape_Sphericity & original_firstorder_90Percentile are continuous variables and were standardized based on the mean and standard deviation of training dataset, Age was Age at diagnosis/ 100, and all other variables are binary)

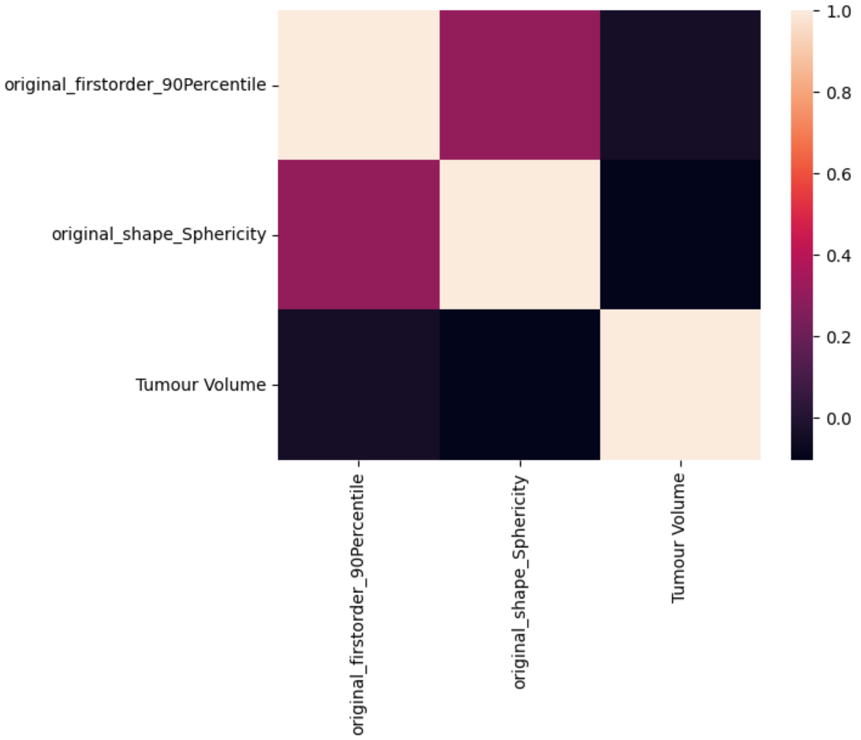


**Figure A1:** Heat map showing the correlation of the selected Radiomic features with Tumour volume
